# Clinical Characteristics of Coronavirus Disease 2019 in Hainan, China

**DOI:** 10.1101/2020.03.19.20038539

**Authors:** Shijiao Yan, Xingyue Song, Feng Lin, Haiyan Zhu, Xiaozhi Wang, Min Li, Jianwen Ruan, Changfeng Lin, Xiaoran Liu, Qiang Wu, Zhiqian Luo, Wenning Fu, Song Chen, Yong Yuan, Shengxing Liu, Jinjian Yao, Chuanzhu Lv

**Author notes:** These authors contributed equally to this work (Shijiao Yan, Xingyue Song, Feng Lin, Haiyan Zhu, Xiaozhi Wang, Min Li, Jianwen Ruan, Changfeng Lin). **Correspondence to:** Jinjian Yao, MD, Department of emergency, Hainan General Hospital, Hainan Affiliated Hospital of Hainan Medical University, Haikou, China. Chuanzhu Lv, MD, Department of Emergency, Hainan Clinical Research Center for Acute and Critical Diseases, The Second Affiliated Hospital of Hainan Medical University, No. 3 Xueyuan Road, Longhua Zone, Haikou 571199, China.

## Abstract

**Background:** Since January 2020, coronavirus disease 2019 (Covid-19) has spread rapidly and developing the pandemic model around the world. Data have been needed on the clinical characteristics of the affected patients in an imported cases as model in island outside Wuhan.

**Methods:** We conducted a retrospective study included all 168 confirmed cases of Covid-19 in Hainan province from 22 January 2020 to 13 March 2020. Cases were confirmed by real-time RT-PCR and were analysed for demographic, clinical, radiological and laboratory data.

**Results:** Of 168 patients, 160 have been discharged, 6 have died and 2 remain hospitalized. The median age was 51.0 years and 51.8% were females. 129 (76.8%) patients were imported cases, and 118 (70.2%), 51 (30.4%) and 52 (31%) of patients lived in Wuhan or traveled to Wuhan, had contact with Covid-19 patients, or had contact with Wuhan residents, respectively. The most common symptoms at onset of illness were fever (65.5%), dry cough (48.8%) and expectoration (32.1%). On admission, ground-glass opacity was the most common radiologic finding on chest computed tomography (60.2%). The elderly people with diabetes, hypertension and CVD are more likely to develop severe cases. Follow-up of 160 discharged patients found that 20 patients (12.5%) had a positive RT-PCR test results of pharyngeal swabs or anal swabs or fecal.

**Conclusions:** In light of the rapid spread of Covid-19 around the world, early diagnosis and quarantine is important to curb the spread of Covid-19 and intensive treatments in early stage is to prevent patients away from critical condition.

## Introduction

In December 2019, patients with pneumonia of unknown causes were found in some medical institutions in Wuhan, Hubei province, China.^1^ On January 7, 2020, the virus was isolated from a sample of a positive patient and the entire genome sequence of the virus was obtained.^2^ Subsequent studies indicated that the virus belonged to the type b coronavirus family, and its genetic characteristics were obviously different from Severe Acute Respiratory Syndrome (SARS) and Middle East Respiratory Syndrome (MERS).^3,4^ The World Health Organization (WHO) declared the new coronavirus pneumonia as a public health emergency of international concern on 31 January 2020, and officially named it “coronavirus disease 2019 (Covid-19)”on February 11, 2020. Covid-19 is mainly transmitted from human-to-human by respiratory droplets and close contact, and may also be transmitted through fecal mouth and aerosol.^5,6^ The clinical manifestations of Covid-19 are very similar to SARS, and patients may develop acute respiratory distress syndrome (ARDS), which is highly likely to require intensive care and even lead to death.^7^ Since January 2020, the Covid-19 has spread rapidly from Wuhan to other parts of China and other countries around the world.^8-10^ As of March 13, 2020, a total of 132,758 laboratory-confirmed cases (80,991 in China) and 4955 deaths (3180 in China) had been reported worldwide.^11^ With the increasing number of confirmed and suspected cases globally, the situation of disease prevention and control is extremely critical.

Since the outbreak of Covid-19, several studies have successively reported the clinical characteristics of patients infected with Covid-19, but almost all the research was conducted in Wuhan or other cities in inland China.^12-15^ Hainan province is an independent island located at the southern tip of China, unlike the rest of China, Hainan province belongs to tropical monsoon climate. Previous studies have found that climate affects the spread of SARS virus.^16^ Therefore, we describe the clinical characteristics and laboratory findings of patients infected with Covid-19 in Hainan province to explore whether it is different from other regions in China, and to provide an insight into the prevention and treatment of Covid-19 across China, especially to other places with similar climate and environment as Hainan province.

## Methods

### Study Design and Participants

We conducted a retrospective study focusing on the clinical characteristics of confirmed cases of Covid-19 in Hainan province from 22 January 2020 (the time of the first confirmed patient in Hainan province) to 14 March 2020. Sputum and throat swab specimens collected from all patients at admission were tested by real time polymerase chain reaction for SARS-Cov-2 RNA. All patients with Covid-19 were diagnosed based on the WHO interim guidance^17^ and the “Diagnosis and Treatment of New Coronavirus Pneumonia (trial version 3)” published by National Health Commission of the People’s Republic of China.^18^ A confirmed case was defined as a positive result to high-throughput sequencing or real-time reverse-transcriptase polymerase-chain-reaction (RT-PCR) assay for nasal and pharyngeal swab specimens.^7^ Only the laboratory-confirmed cases were included in the analysis.

The study was approved by the institutional ethics board of the Second Affiliated Hospital of Hainan Medical University (No. 2020R003). Written informed consent was waived due to the urgent need to collect clinical data on this emerging disease.

### Data Collection

All medical records of patients were extracted by a doctor assigned by local hospital and sent to the data processing center in Haikou. A team of experienced respiratory clinicians reviewed and abstracted the data. Data were entered into a computerized database and cross-checked. If the data were with mission or doubt, the coordinators in Haikou contacted the doctor who is responsible for the treatment of the patient for clarification.

The epidemiological characteristics (demographic data, exposure history, and underlying comorbidities), clinical (symptoms and signs), laboratory (ie, complete blood count, blood chemistry, coagulation test, liver and renal function, electrolytes) and radiological findings (chest X-ray or computed tomography), treatment (ie, antiviral therapy, corticosteroid therapy, respiratory support) and outcomes (remained in hospital, discharges, death) data were extracted from electronic medical records. The severity of Covid-2019 was defined based on the “Diagnosis and Treatment of New Coronavirus Pneumonia (trial version 7)” published by National Health Commission of the People’s Republic of China.^19^ Severe patients are defined as having been critically ill during their hospitalization. Fever was defined as an axillary temperature of 37.3°C or higher. Acute respiratory distress syndrome (ARDS) was defined according to the Berlin definition.^20^ Acute kidney injury was identified according to the Kidney Disease: Improving Global Outcomes definition.^21^ Cardiac injury was defined if the serum levels of cardiac biomarkers (eg, troponin I) were above the 99th percentile upper reference limit or new abnormalities were shown in electrocardiography and echocardiography.^22^ The durations from onset of disease to first hospital visit, admission, severe, discharge or death were recorded. The clinical outcomes were monitored up to March 13, 2020, the final date of follow-up.

### Statistical Analysis

Categorical variables were presented as the counts and percentages in each category, and chi-square tests and Fisher’s exact tests were used for comparing as appropriate. Continuous variables were presented as medians and interquartile ranges (IQR) and compared with Wilcoxon rank-sum tests. All analyses were performed using SPSS (Statistical Package for the Social Sciences) version 22.0 software (SPSS Inc), and the statistical significance was two-sided *P* values <0.05. Distribution map was plotted using ArcGis version 10.2.2

## Results

As of March 13, 2020, a total of 168 patients (Non-severe: 132; Severe: 36) were infected with Covid-19 in Hainan province. The distribution of patients infected with Covid-19 across Hainan province is presented in Figure 1. The demographic and clinical characteristics of patients by severity group are presented in Table 1. The median age was 51.0 years (IQR, 36-62), 51.8% were females and 33.3% were the retired. 129 (76.8%) patients were imported cases, and 118 (70.2%), 51 (30.4%) and 52 (31%) of the patients lived in Wuhan or traveled to Wuhan, had contact with Covid-19 patients, or had contact with Wuhan residents, respectively.

**Table 1.** **Demographic and Clinical Characteristics of Patients Infected with Coronavirus Disease 2019 in Hainan Province**

| **Characteristics** | **All patients**<br>**( N=168 )** | **Non-severe**<br>**( N=132 )** | **Severe**<br>**( N=36 )** | ***P*** |
| --- | --- | --- | --- | --- |
| **Age** |  |  |  |  |
| **Median (IQR)-yrs** | 51 (36-62) | 49 (34-60) | 61 (50.3-68) | **0.002** |
| **Distributions - no. (%)** |  |  |  | **0.009** |
| 0-17 yr | 8 (4.8) | 8 (6.1) | 0 (0) |  |
| 18-49 yr | 68 (40.5) | 60 (45.5) | 8 (22.2) |  |
| 50-64 yr | 59 (35.1) | 43 (32.6) | 16 (44.4) |  |
| ≥65 yr | 33 (19.6) | 21 (15.9) | 12 (33.3) |  |
| **Sex - no. (%)** |  |  |  | 0.17 |
| Male | 81 (48.2) | 60 (45.5) | 21 (58.3) |  |
| Female | 87 (51.8) | 72 (54.5) | 15 (41.7) |  |
| **Job - no. (%)** |  |  |  | **0.01** |
| Retired | 56 (33.3) | 37 (28) | 19 (52.8) |  |
| Medical staff | 1 (0.6) | 0 (0) | 1 (2.8) |  |
| Worker/Farmer | 23 (13.7) | 21 (15.9) | 2 (5.6) |  |
| Service staff | 37 (22) | 33 (25) | 4 (11.1) |  |
| Others | 51 (30.4) | 41 (31.1) | 10 (27.8) |  |
| **Sources of cases - no. (%)** |  |  |  | 0.87 |
| Imported | 129 (76.8) | 101 (76.5) | 28 (77.8) |  |
| Local | 39 (23.2) | 31 (23.5) | 8 (21.9) |  |
| **Exposure to source of transmission** |  |  |  |  |
| **within past 14 days - no. (%)** |  |  |  |  |
| Living in Wuhan or travel in Wuhan | 118 (70.2) | 93 (70.5) | 25 (69.4) | 0.91 |
| Had contact with Covid-19 patients | 51 (30.4) | 42 (31.8) | 9 (25) | 0.43 |
| Had contact with Wuhan residents | 52 (31) | 38 (28.8) | 14 (38.9) | 0.25 |
| Aggregation disease | 102 (60.7) | 82 (62.1) | 20 (55.6) | 0.48 |
| **Symptoms - no. (%)** |  |  |  |  |
| Dry cough | 82 (48.8) | 63 (47.7) | 19 (52.8) | 0.59 |
| Expectoration | 54 (32.1) | 43 (32.6) | 11 (30.6) | 0.82 |
| Nasal congestion | 10 (6) | 6 (4.5) | 4 (11.1) | 0.28 |
| Sore throat | 19 (11.3) | 13 (9.8) | 6 (16.7) | 0.40 |
| Rhinorrhoea | 6 (3.6) | 6 (4.5) | 0 (0) | 0.43 |
| Diarrhea | 12 (7.1) | 8 (6.1) | 4 (11.1) | 0.5 |
| Abdominal pain | 7 (4.2) | 5 (3.8) | 2 (5.6) | 1.00 |
| Myalgia | 14 (8.3) | 11 (8.3) | 3 (8.3) | 1.00 |
| Nausea | 9 (5.4) | 6 (4.5) | 3 (8.3) | 0.63 |
| Vomiting | 7 (4.2) | 3 (2.3) | 4 (11.1) | 0.06 |
| Anorexia | 14 (8.3) | 8 (6.1) | 6 (16.7) | 0.09 |
| Dizziness | 12 (7.1) | 8 (6.1) | 4 (11.1) | 0.50 |
| Headache | 14 (8.3) | 9 (6.8) | 5 (13.9) | 0.31 |
| Weakness | 47 (28) | 33 (25) | 14 (38.9) | 0.10 |
| Dyspnea | 7 (4.2) | 2 (1.5) | 5 (13.9) | **0.005** |
| Arthralgia | 2 (1.2) | 2 (1.5) | 0 (0) | 0.62 |
| Anhelation | 22 (13.1) | 15 (11.4) | 7 (19.4) | 0.32 |
| chest tightness | 24 (14.3) | 17 (12.9) | 7 (19.4) | 0.32 |
| Pharyngeal congestion | 1 (0.6) | 1 (0.8) | 0 (0) | 0.79 |
| Pharyngeal discomfort | 7 (4.2) | 5 (3.8) | 2 (5.6) | 1.00 |
| Dry throat | 5 (3) | 3 (2.3) | 2 (5.6) | 0.64 |
| Chill | 10 (6) | 8 (6.1) | 2 (5.6) | 1.00 |
| Shortness of breath | 4 (2.4) | 3 (2.3) | 1 (2.8) | 0.62 |
| Hemoptysis | 1 (0.6) | 0 (0) | 1 (2.8) | 0.21 |
| **Fever on admission** |  |  |  |  |
| **Median temperature (IQR) — °C** | 36.8 (36.5-37.3) | 36.8 (36.5-37.3) | 37.0 (36.7-37.5) | **0.04** |
| **Patients - no. (%)** | 110 (65.5) | 82 (62.1) | 28 (77.8) | 0.08 |
| **Distribution of temperature - no. (%)** |  |  |  | 0.15 |
| <37.5°C | 131 (78) | 106 (80.3) | 25 (69.4) |  |
| 37.5-38.0 °C | 24 (14.3) | 18 (13.6) | 6 (16.7) |  |
| 38.1-39.0°C | 12 (7.1) | 8 (6.1) | 4 (11.1) |  |
| >39.0°C | 1 (0.6) | 0 (0) | 1 (2.8) |  |
| **Fever during hospitalization** |  |  |  |  |
| **Patients - no. (%)** | 97 (57.7) | 67 (50.8) | 30 (87.5) | **<0.001** |
| **Distribution of temperature - no./total no. (%)** |  |  |  | **0.005** |
| <37.5°C | 11/97 (11.3) | 8/67 (11.9) | 3/30 (10) |  |
| 37.5-38.0 °C | 38/97 (39.2) | 30/67 (44.8) | 8/30 (26.7) |  |
| 38.1-39.0°C | 36/97 (37.1) | 26/67 (38.8) | 10/30 (33.3) |  |
| >39.0°C | 12/97 (12.4) | 3/67 (4.5) | 9/30 (30) |  |
| **Respiratory rate, median (IQR)** | 20 (20-20) | 20 (20-20) | 20 (20-22.8) | **0.58** |
| **Heart rate, median (IQR), bpm** | 82.5 (76-92) | 80.5 (75-88) | 92 (82.3-102.3) | **<0.001** |
| **Systolic Blood Pressure, mmHg** | 124 (115-136.3) | 123.5 (115.8-135) | 126 (112-139.5) | 0.80 |
| **Diastolic blood pressure, mmHg** | 78 (72-86) | 78.5 (72-86) | 76 (73-83.5) | 0.63 |
| **Comorbidities - no. (%)** |  |  |  |  |
| Hypertension | 24 (14.3) | 13 (9.8) | 11 (30.6) | **0.002** |
| Diabetes | 12 (7.1) | 5 (3.8) | 7 (19.4) | **0.004** |
| Cardiovascular disease | 12 (7.1) | 6 (4.5) | 6 (16.7) | **0.03** |
| Cancer | 2 (1.2) | 2 (1.5) | 0 (0) | 0.62 |
| Chronic kidney disease | 1 (0.6) | 0 (0) | 1 (2.8) | 0.21 |
| Chronic liver disease | 6 (3.6) | 3 (2.3) | 3 (8.3) | 0.22 |
| Lung disease | 10 (6) | 6 (4.5) | 4 (11.1) | 0.28 |
| Chronic bronchitis/asthma /other |  |  |  |  |
| upper respiratory diseases | 2 (1.2) | 2 (1.5) | 0 (0) | 0.62 |
| Thyroid disease | 2 (1.2) | 1 (0.8) | 1 (2.8) | 0.38 |
| Lithiasis | 2 (1.2) | 1 (0.8) | 1 (2.8) | 0.38 |

**Figure 1.**
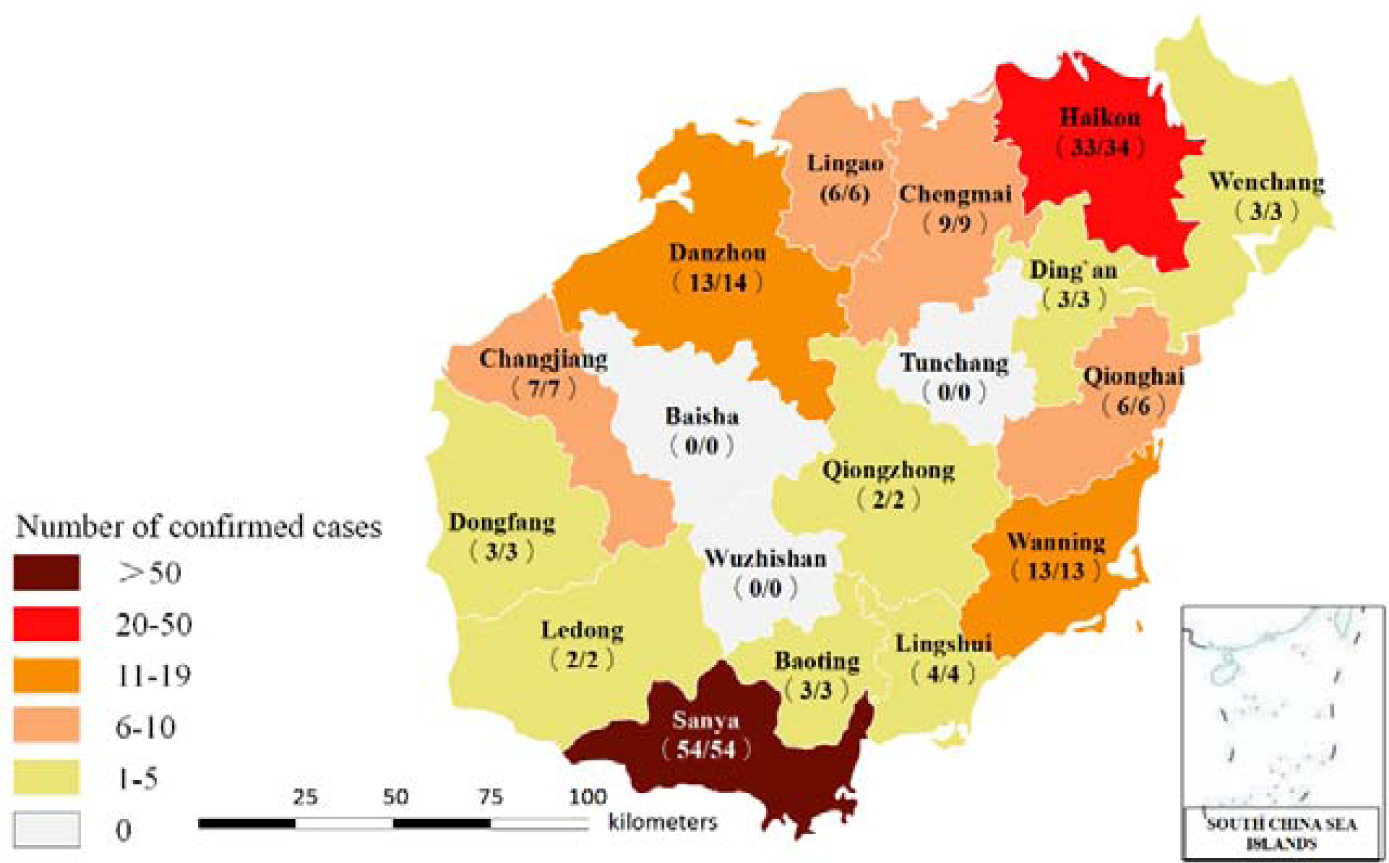
Distribution of Patients infected with Coronavirus Disease 2019 across Hainan Province. The numerator denotes the number of discharge patients and the denominator denotes the number of laboratory-confirmed cases

The most common symptoms at onset of illness were fever (65.5%), dry cough (48.8%) and expectoration (32.1%), while rhinorrhea (3.6%), abdominal pain (4.2%), vomiting (4.2%), dyspnea (4.2%), arthralgia (1.2%), pharyngeal congestion (0.6%), pharyngeal discomfort (4.2%), dry throat (3.0%), shortness of breath (2.4%) and hemoptysis (0.6%) were rare. On admission, 24 (14.3%), 12 (7.1%), and 12 (7.1%) patients are with history of hypertension, diabetes, and cardiovascular disease, respectively.

Compared with non-severe patients, severe patients were significantly older (median, 61 years vs 49 years; *P* < 0.05), were more likely to have higher temperature (median, 37.0**°**C vs 36.8**°**C; *P* < 0.05) and heart rate (median, 92.0 bpm vs 80.5 bpm; *P* < 0.05) on admission, and more likely to have underlying comorbidities, including hypertension (30.6% vs 9.8%; *P* < 0.05), diabetes (19.4% vs 3.8%; *P* < 0.05) and cardiovascular disease (CVD) (16.7% vs 4.5%; *P* < 0.05).

### Radiologic and Laboratory Findings

The radiologic and laboratory findings on admission are shown in Table 2. Of 133 patients who underwent chest computed tomography (CT), the most common patterns were ground-glass opacity (60.2%) and bilateral patchy shadowing (42.9%). Of 51 patients who underwent chest X-ray, the most common patterns were bilateral patchy shadowing (35.3%) and local patchy shadowing (31.4%). Typical chest CT findings of infected patients on admission were shown in Figure 2.

**Table 2.** Radiographic and Laboratory Findings of Patients Infected with Coronavirus Disease 2019 in Hainan Province.

|  | Normal<br>Range |  | All patients |  | Non-severe |  | Severe | *P* |
| --- | --- | --- | --- | --- | --- | --- | --- | --- |
| Radiologic findings - no. (%) |  | N |  | N |  | N |  |  |
| Abnormalities on chest CT |  |  |  |  |  |  |  |  |
| Ground-glass opacity | N/A | 133 | 80 (60.2) |  | 67/116 (57.8) |  | 13/17 (76.5) | 0.14 |
| Local patchy shadowing | N/A | 133 | 29 (21.8) |  | 29/116 (25) |  | 0/17 (0) | **0.04** |
| Bilateral patchy shadowing | N/A | 133 | 57 (42.9) |  | 46/116 (39.7) |  | 11/17 (64.7) | 0.05 |
| Interstitial abnormalities | N/A | 133 | 29 (21.8) |  | 26/116 (22.4) |  | 3/17 (17.6) | 0.9 |
| Abnormalities on chest X-ray |  |  |  |  |  |  |  |  |
| Ground-glass opacity | N/A | 51 | 7 (13.7) |  | 2/29 (6.9) |  | 5/22 (22.7) | 0.22 |
| Local patchy shadowing | N/A | 51 | 16 (31.4) |  | 11/29 (37.9) |  | 5/22 (22.7) | 0.25 |
| Bilateral patchy shadowing | N/A | 51 | 18 (35.3) |  | 8/29 (27.6) |  | 10/22 (45.5) | 0.19 |
| Interstitial abnormalities | N/A | 51 | 4 (7.8) |  | 0/29 (0) |  | 4/22 (18.2) | 0.06 |
| **Laboratory findings** |  |  |  |  |  |  |  |  |
| White blood cell count, $\times 10^9/L$ | 3.5-9.5 | 167 | 4.8 (4.0-6.3) | 132 | 4.7 (3.9-5.9) | 35 | 5.6 (4.3-7.8) | **0.006** |
| Increased - no. (%) |  | 167 | 8 (4.8) | 132 | 2 (1.5) | 35 | 6 (21.9) |  |
| Decreased - no. (%) |  | 167 | 40 (24) | 132 | 36 (27.3) | 35 | 4 (21.9) |  |
| Platelet count, $\times 10^9/L$ | 125-350 | 167 | 189 (147-249) | 132 | 200.5 (152-253) | 35 | 152 (117-210) | **0.005** |
| Increased - no. (%) |  | 167 | 17 (10.2) | 132 | 15 (11.4) | 35 | 2 (21.9) |  |
| Decreased - no. (%) |  | 167 | 10 (6) | 132 | 5 (3.8) | 35 | 5 (21.9) |  |
| Lymphocyte ratio, % | 20-50 | 166 | 26.2 (17.2-31.8) | 131 | 27.9 (20.8-34.5) | 35 | 16.8 (8.2-21.5) | **<0.001** |
| Neutrophil ratio, % | 40-75 | 167 | 64 (57.8-73.1) | 132 | 62.1 (55.6-69.2) | 35 | 74.8 (67.6-83.1) | **<0.001** |
| Lymphocyte count, $\times 10^9/L$ | 1.1-3.2 | 165 | 1.2 (0.8-1.5) | 130 | 1.3 (0.9-1.6) | 35 | 0.8 (0.6-1.2) | **<0.001** |
| Increased - no. (%) |  | 165 | 3 (1.8) | 130 | 3 (2.3) | 35 | 0 (0) |  |
| Decreased - no. (%) |  | 165 | 72 (43.6) | 130 | 49 (37.7) | 35 | 23 (65.7) |  |
| Monocyte count, $\times 10^9/L$ | 0.1-0.6 | 165 | 0.4 (0.3-0.5) | 130 | 0.4 (0.3-0.5) | 35 | 0.4 (0.3-0.6) | 0.49 |
| Neutrophil count, $\times 10^9/L$ | 1.8-6.3 | 166 | 3.1 (2.3-4.3) | 131 | 2.9 (2.2-3.9) | 35 | 4.2 (3.0-7.7) | **<0.001** |
| Increased - no. (%) |  | 166 | 10 (6) | 131 | 3 (2.3) | 35 | 7 (21.9) |  |
| Decreased - no. (%) |  | 166 | 19 (11.4) | 131 | 16 (12.2) | 35 | 3 (21.9) |  |
| Eosinophil count, $\times 10^9/L$ | 0.02-0.52 | 165 | 0.03 (0-0.1) | 130 | 0.04 (0.01-0.1) | 35 | 0 (0-0.02) | **<0.001** |
| C-reactive protein, mg/L | 0-8.2 | 97 | 17 (3.8-35.9) | 69 | 11.8 (1.2-26.2) | 28 | 33.8 (18.5-64.4) | **<0.001** |
| Increased - no. (%) |  | 97 | 65 (67) | 69 | 40 (58) | 28 | 25 (21.9) |  |
| Prothrombin time, s | 9.8-13.2 | 95 | 11.6 (11-12.3) | 69 | 11.6 (11-12.2) | 26 | 11.7 (11.2-12.7) | 0.22 |
| Increased - no. (%) |  | 95 | 10 (10.5) | 69 | 5 (7.2) | 26 | 5 (21.9) |  |
| Activated partial thromboplastin<br>time, s | 21-36 | 96 | 29.1 (26.4-32.5) | 69 | 29.2 (26.6-32.7) | 27 | 28.9 (25.3-31.8) | 0.69 |
| Increased - no. (%) |  | 96 | 8 (8.3) | 69 | 4 (5.8) | 27 | 4 (21.9) |  |
| D-dimer, mg/L | <0.5 | 46 | 0.3 (0.2-0.7) | 31 | 0.3 (0.2-0.5) | 15 | 0.6 (0.3-2.6) | **0.003** |
| Increased - no. (%) |  | 46 | 14 (30.4) | 31 | 7 (22.6) | 15 | 7 (46.7) |  |
| Potassium, mmol/L | 3.5-5.3 | 124 | 3.8 (3.5-4.1) | 93 | 3.8 (3.5-4.1) | 31 | 3.7 (3.4-4) | 0.64 |
| Sodium, mmol/L | 137-147 | 124 | 138.8 (136-142.2) | 93 | 139 (136.5-142.3) | 31 | 137 (132-142) | 0.12 |
| Chloride, mmol/L | 99-110 | 123 | 104.1 (102-106.4) | 92 | 104.8 (103-106.6) | 31 | 103 (100-106) | **0.02** |
| Calcium, mmol/L | 2.2-2.7 | 118 | 2.3 (2.2-2.4) | 88 | 2.3 (2.2-2.4) | 30 | 2.2 (2.1-2.3) | **0.03** |
| Phosphorus, mmol/L | 0.85-1.51 | 95 | 1.02 (0.84-1.32) | 69 | 1.05 (0.88-1.35) | 26 | 0.92 (0.79-1.12) | 0.06 |
| Magnesium, mmol/L | 0.75-1.02 | 101 | 0.87 (0.79-0.96) | 75 | 0.87 (0.79-0.97) | 26 | 0.86 (0.79-0.93) | 0.53 |
| Blood urea nitrogen, mmol/L | 3.6-9.5 | 122 | 3.7 (2.9-4.5) | 93 | 3.6 (2.9-4.3) | 29 | 4.1 (2.9-7.0) | 0.05 |
| Increased - no. (%) |  | 122 | 4 (3.3) | 93 | 0 (0) | 29 | 4 (21.9) |  |
| Decreased - no. (%) |  | 122 | 24 (19.7) | 93 | 19 (20.4) | 29 | 5 (21.9) |  |
| Creatinine, µmol/L | 57-111 | 118 | 62 (49-75.1) | 88 | 62.1 (49.8-72.0) | 30 | 57.3 (48.6-87.1) | 0.69 |
| Increased - no. (%) |  | 118 | 4 (3.4) | 88 | 0 (0) | 30 | 4 (13.3) |  |
| Decreased - no. (%) |  | 118 | 8 (6.8) | 88 | 5 (5.7) | 30 | 3 (10) |  |
| Total protein, g/L | 65-85 | 113 | 65 (60.2-68.6) | 83 | 66 (60.7-69.2) | 30 | 62.3 (57.3-67.0) | **0.01** |
| Decreased - no. (%) |  | 113 | 49 (43.4) | 83 | 30 (36.1) |  | 19 (63.3) |  |
| Albumin, g/L | 40-55 | 115 | 41.7 (38.8-45) | 84 | 42.4 (39.9-47.1) | 31 | 40.4 (33.2-42.2) | **0.001** |
| Increased - no. (%) |  | 115 | 3 (2.6) | 84 | 2 (2.4) | 31 | 1 (3.2) |  |
| Decreased - no. (%) |  | 115 | 32 (27.8) | 84 | 18 (21.4) | 31 | 14 (45.2) |  |
| Alanine aminotransferase, U/L | 9-50 | 112 | 22.6 (16-35) | 81 | 20.9 (14.9-33) | 31 | 29 (16.6-40.8) | 0.08 |
| Increased - no. (%) |  | 112 | 9 (8) | 81 | 5 (6.2) | 31 | 4 (12.9) |  |
| Decreased - no. (%) |  | 112 | 6 (5.4) | 81 | 4 (4.9) | 31 | 2 (6.5) |  |
| Aspartate aminotransferase, U/L | 15-40 | 104 | 25.3 (20-38.63) | 75 | 24.3 (20-31) | 29 | 37.1 (24-51.5) | **0.004** |
| Increased - no. (%) |  | 104 | 18 (17.3) | 75 | 7 (9.3) | 29 | 11 (37.9) |  |
| Decreased - no. (%) |  | 104 | 2 (1.9) | 75 | 1 (1.3) | 29 | 1 (3.4) |  |
| Lactate dehydrogenase, U/L | 120-250 | 93 | 207 (169.5-274.5) | 67 | 188 (164-248) | 26 | 256.5 (211.3-413.8) | **0.001** |
| Total bilirubin, mmol/L | 0-26 | 112 | 8.8 (6.3-13.4) | 81 | 8.2 (5.7-10.6) | 31 | 10.5 (8.4-14.6) | **0.004** |
| Creatine kinase, U/L | 50-310 | 109 | 68 (44.5-113.5) | 80 | 68 (47-107) | 29 | 75 (34-144) | 0.92 |
| Increased - no. (%) |  | 109 | 7 (6.4) | 80 | 3 (3.8) | 29 | 4 (13.8) |  |
| Decreased - no. (%) |  | 109 | 9 (8.3) | 80 | 7 (8.8) | 29 | 2 (6.9) |  |
| Creatine kinase-MB, U/L | 0-24 | 109 | 12 (8-15.8) | 80 | 12.1 (9.0-15.7) | 29 | 9.9 (6.5-18.3) | 0.35 |
| Procalcitonin, ng/mL | <0.046 | 103 | 0.04 (0.026-0.069) | 81 | 0.038 (0.02-0.06) | 22 | 0.067 (0.032-0.257) | **0.007** |
| Increased - no. (%) |  | 103 | 26 (25.2) | 81 | 19 (23.5) | 22 | 7 (31.8) |  |

Data are presented as frequency (percentage) and median (IQR) unless otherwise indicated. Increased means over the upper limit of the normal range and decreased means below the lower limit of the normal range.

**Figure 2.**
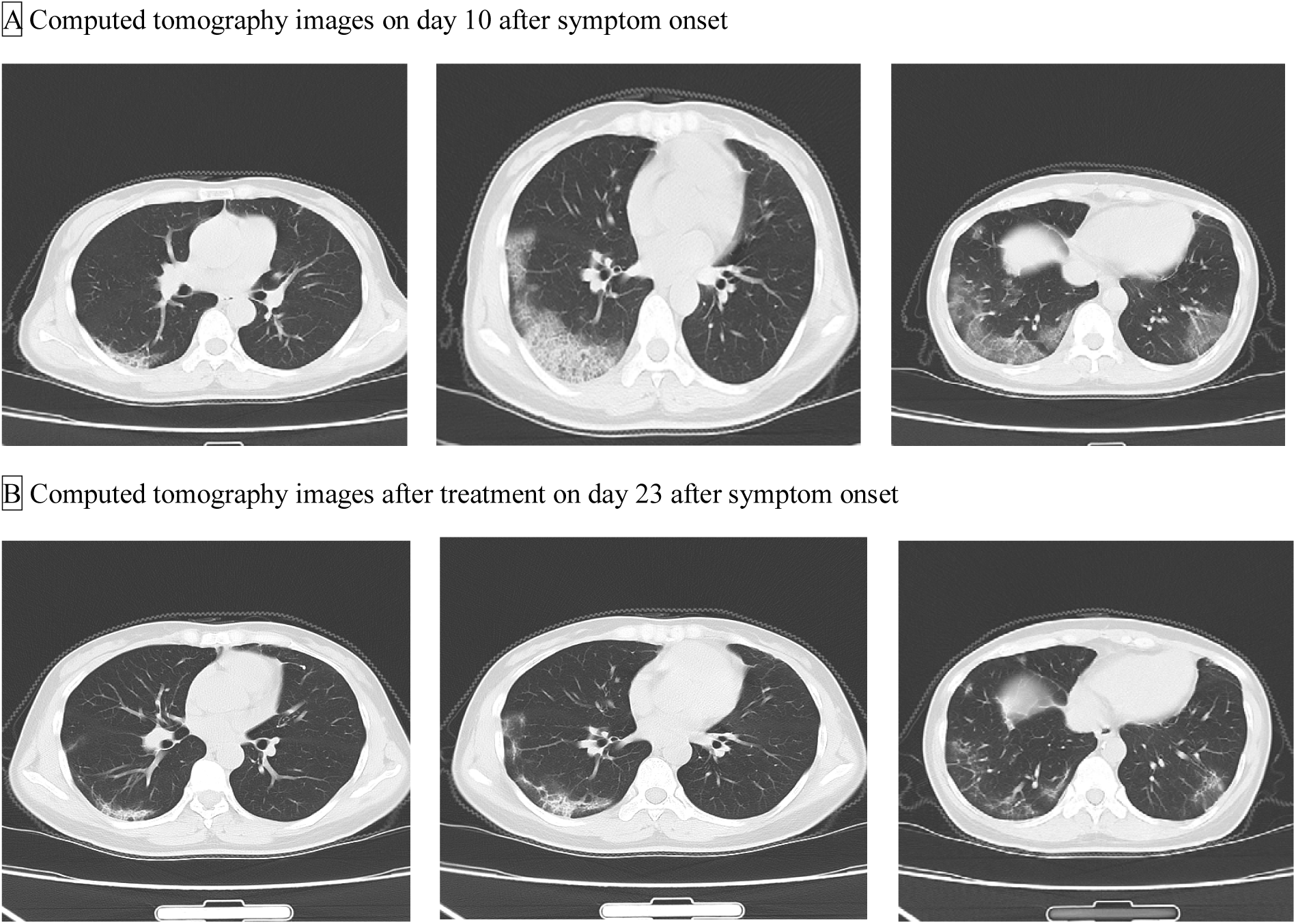
Chest Computed Tomographic Images of a 50-Year-Old Severe Patient Infected With Coronavirus Disease 2019. A, Chest computed tomographic images obtained on February 10, 2020, show ground glass opacity in both lungs on day 10 after symptom onset. B, Images taken on February 23, 2020, show fibrous cord shadows in both lungs, and the absorption of inflammation.

On admission, the levels of lymphocyte, white blood cell and neutrophil were decreased in 72 (72%) of 165 patients, 40 (24%) of 167 patients and 19 (11.4%) patients, respectively, while the levels of C-reactive protein, D-dimer, prothrombin time and activated partial thromboplastin time were increased in 65 (67%) of 97 patients, 14 (30.4%) of 46 patients, 10 (10.5%) of 95 patients and 8 (8.3%) of 96 patients, respectively. There were numerous differences in laboratory findings between non-severe patients and severe patients, including white blood cell count, platelet count, lymphocyte ratio, neutrophil ratio, lymphocyte count, neutrophil count, eosinophil count, C-reactive protein, D-dimer, chloride, calcium, total protein, albumin, aspartate aminotransferase, lactate dehydrogenase, total bilirubin and procalcitonin (All *P* <0.05). The dynamic changes in 6 clinical laboratory parameters of 36 severe patients, including hematological and biochemical parameters were shown in Supplemental Figure 1.

### Clinical Outcomes, Complications and Treatment

Of the all 168 patients, 160 have been discharged, 6 have died and 2 remain hospitalized (Table 3). The median time from onset of symptoms to first hospital visit, diagnosis and hospital admission was 1 day (IQR, 0-4), 4.5 days (IQR, 2-7) and 5 days (IQR, 2-7), respectively (Table 4). For severe patients, the median time from onset of symptoms to severe illness was 11 days (IQR, 7-13). For discharged patients, the median time from onset of symptoms to discharged was 22 days (IQR, 17-27), and the median time from hospital admission to discharged was 17 days (IQR, 12-21). For dead patients, the median time from onset of symptoms to death was 17 days (IQR, 10.5-37.3), and the median time from hospital admission to death was 13 days (IQR, 1.5-28.3). Follow-up of 160 discharged patients found that 20 patients (12.5%) had a positive RT-PCR test results of pharyngeal swabs or anal swabs or faeces.

**Table 3.** **Complications, Treatments, and Clinical Outcomes of Patients Infected with Coronavirus Disease 2019 in Hainan Province**

| **Variable** | **All patients**<br>**( N=168 )** | **Non-severe**<br>**( N=132 )** | **Severe**<br>**( N=36 )** | ***P*** |
| --- | --- | --- | --- | --- |
| **Clinical outcome- no. (%)** |  |  |  | **<0.001** |
| Discharged | 160 (95.2) | 130 (98.5) | 30 (83.3) |  |
| Died | 6 (3.6) | 0 (0) | 6 (16.7) |  |
| Remained in hospital | 2 (1.2) | 2 (1.5) | 2 (5.6) |  |
| **Course- Median (IQR), days** |  |  |  |  |
| Time from onset of symptoms to first<br>hospital visit (N=168) | 1 (0-4) | 1 (0-4) | 2 (0-4.8) | **<0.001** |
| Time from onset of symptoms to<br>diagnosis (N=168) | 4.5 (2-7) | 4 (1.3-7) | 6 (2.3-11.5) | **<0.001** |
| Time from onset of symptoms to<br>hospital admission (N=168) | 5 (2-7) | 4.5 (2-7) | 7 (3-10) | **<0.001** |
| Time from onset of symptoms to severe<br>illness (N=36) | 11 (7-13) | N/A | N/A | N/A |
| Time from onset of symptoms to<br>discharge (N=160) | 22 (17-27) | N/A | N/A | N/A |
| Time from hospital admission to<br>discharge (N=160) | 17 (12-21) | N/A | N/A | N/A |
| Time from onset of symptoms onset to | 17 (10.5-37.3) | N/A | N/A | N/A |
| death (N=6) |  |  |  |  |
| Time from hospital admission to death<br>(N=6) | 13 (1.5-28.3) | N/A | N/A | N/A |
| **Positive RT-PCR test after discharge-**<br>**no./total no. (%)** | 20/160 (12.5) | 18/130 (13.8) | 2/30 (6.7) | 0.44 |
| **Complications- no. (%)** |  |  |  |  |
| Acute cardiac injury | 4 (2.4) | 0 (0) | 4 (11.1) | **0.002** |
| Acute respiratory distress syndrome | 17 (10.1) | 1 (0.8) | 16 (44.4) | **<0.001** |
| Acute kidney injury | 6 (3.6) | 0 (0) | 3 (8.3) | **<0.001** |
| Sepsis | 5 (3) | 0 (0) | 5 (13.9) | **<0.001** |
| Acute lung injury | 4 (2.4) | 0 (0) | 4 (11.1) | **0.002** |
| Secondary bacterial pneumonia | 12 (7.1) | 2 (1.5) | 7 (19.4) | **<0.001** |
| Arrhythmia | 2 (1.2) | 0 (0) | 1 (2.8) | **0.04** |
| Epilepsy | 2 (1.2) | 0 (0) | 2 (5.6) | **0.04** |
| Shock | 7 (4.2) | 0 (0) | 7 (19.4) | **<0.001** |
| Secondary fungal infection | 2 (1.2) | 0 (0) | 1 (2.8) | **0.04** |
| Multiple organ dysfunction | 2 (1.2) | 0 (0) | 2 (5.6) | **0.04** |
| Respiratory failure | 10 (6) | 0 (0) | 3 (8.3) | **<0.001** |
| Disseminated intravascular coagulation | 2 (1.2) | 0 (0) | 1 (2.8) | **0.04** |
| Gastrointestinal flora disorder | 1 (0.6) | 0 (0) | 1 (2.8) | 0.22 |
| Hypoproteinemia | 3 (1.8) | 0 (0) | 3 (8.3) | **0.009** |
| Electrolyte disturbances | 3 (1.8) | 0 (0) | 3 (8.3) | **0.009** |
| Acute liver injury | 4 (2.4) | 0 (0) | 4 (11.1) | **0.002** |
| Digestive tract hemorrhage | 4 (2.4) | 0 (0) | 4 (11.1) | **0.002** |
| Pneumothorax | 2 (1.2) | 0 (0) | 2 (5.6) | **0.04** |
| Metabolic disorder | 2 (1.2) | 0 (0) | 2 (5.6) | **0.04** |
| **Treatments- no. (%)** |  |  |  |  |
| Nasal cannula oxygen | 97 (57.7) | 70 (53) | 27 (75) | **0.02** |
| Non-invasive ventilator assisted breathing | 13 (7.7) | 2 (1.5) | 11 (30.6) | **<0.001** |
| Trachea cannula | 10 (6) | 0 (0) | 10 (27.8) | **<0.001** |
| Tracheotomy | 2 (1.2) | 0 (0) | 2 (5.6) | **0.04** |
| Mask oxygen inhalation | 3 (1.8) | 0 (0) | 3 (8.3) | **0.009** |
| Cardio-pulmonary resuscitation | 5 (3) | 0 (0) | 5 (13.9) | **<0.001** |
| Antibiotics therapy | 119 (70.8) | 87 (65.9) | 32 (88.9) | **0.007** |
| Antiviral therapy | 155 (92.3) | 120 (90.9) | 35 (97.2) | 0.37 |
| Antifungal therapy | 8 (4.8) | 2 (1.5) | 6 (16.7) | **0.001** |
| Immunopotentiator | 91 (54.2) | 65 (49.2) | 26 (72.2) | **0.01** |
| Glucocorticoid | 27 (16.1) | 8 (6.1) | 19 (52.8) | **<0.001** |
| Acid suppression drugs | 32 (19) | 16 (12.1) | 16 (44.4) | **<0.001** |
| Expectorant and antitussive drugs | 22 (13.1) | 10 (7.6) | 12 (33.3) | **<0.001** |
| Intestinal flora regulating drugs | 71 (42.3) | 52 (39.4) | 19 (52.8) | 0.15 |
| Gastric mucosa repair agent | 17 (10.1) | 10 (7.6) | 7 (19.4) | 0.08 |
| Immunosuppressor | 2 (1.2) | 0 (0) | 2 (5.6) | **0.04** |
| Pseudoephedrine | 2 (1.2) | 0 (0) | 2 (5.6) | **0.04** |
| M-receptor inhibitors | 1 (0.6) | 0 (0) | 1 (2.8) | 0.22 |
| Nitrates | 1 (0.6) | 0 (0) | 1 (2.8) | 0.22 |
| Ca<sup>2+</sup>-receptor blocker | 7 (4.2) | 1 (0.8) | 6 (16.7) | **<0.001** |
| β-blocker | 3 (1.8) | 3 (2.3) | 0 (0) | 0.48 |
| Traditional Chinese medicine | 30 (17.9) | 25 (18.9) | 5 (13.9) | 0.40 |

During hospital admission, the common complication was ARDS (10.1%), secondary bacterial pneumonia (7.1%) and shock (4.2%). Compared with non-severe patients, severe patients are more prone to almost all complications. During the hospital admission, 97 (57.7%) and 13 (7.7%) patients received nasal catheter oxygen and non-invasive ventilator assisted breathing, respectively. Trachea cannula and tracheotomy were performed in 10 and 2 severe patients, respectively. 155 (92.3%) patients received antiviral treatment, 119 (70.8%) were given antibiotic treatment, 91 (54.2%) were given immunopotentiator, 71 (42.3%) patients received intestinal flora regulating drugs, and 27 (16.1%) were given glucocorticoid.

## Discussion

Hainan province as a tropical island in southern China, the aged from the north and middle part of China prefer to spend their winter in Hainan province due to the warmest winter in China at end of every year, especially a part of them has pulmonary, cardiac, metabolic disease. The conditions shaped the epidemic and morbidity of Covid-19 and the demographics and clinical profile of Covid-19 patients distinctive from other places of China. To our knowledge, we first exemplify the Covid-19 outbreak of a real island scenario in an imported Covid-19 epidemic model for worldwide. To date, there are 168 confirmed cases in Hainan province, including 129 (76.8%) imported cases. 36 (21.4%) patients developed into severe illness after admission to the designated hospital, and fatality rate was 3.6%.

The rate of severe illness and death in our study were higher than a study conducted in the multicenter^23^ and a study conducted in Wenzhou, Zhejiang province,^12^ but lower than studies conducted in Wuhan.^7,14,15^ The reason may be that a large number of elderly people come to Hainan at end of each year for avoiding cold winter. 55 (32.7%) patients over 60 years old in our study, this group may have more comorbidities, and more likely to develop into severe illness once infected with Covid-19. Consistent with previous findings,^15,23^ we found that the older and patients with comorbidities (hypertension, diabetes and CVD) were more likely to develop severe illness. All death in patients (5 imported cases and 1 local case) have a kind of comorbidity, 2 of them have two comorbidities. The possible reason may be the chronic course of hypertension and diabetes leading consistent damage to the micro-vascular walls, resulting in both inefficacy of immune cell infiltration and drug deliver into lung areas.^24,25^ In our study, 8 patients were young patients aged 0 to 17 years, the youngest was 3 months and 19 days, and the oldest was 17 years old. The 8 adolescent patients were all relatively mild without significant changes in radiology and laboratory test, moreover, none of them progressed to critical ill case, similar results have also reported in another study conducted in Guangzhou.^26^ The mechanism responsible for this phenomenon is not clear yet and further research is needed to explore the possible causes.

In consistence with previous studies, fever, dry cough and expectoration were the most common symptoms in our study, and it’s worth noting that not all patients infected with Covid-19 had fever, or fever at initial stage.^23,27^ Fever, as an alarm syndrome, is less frequent in Covid-19 than other coronavirus (MERS and SARS), so more attention should be paid to Covid-19 patients without fever for their role as source of infection. Moreover, vomit and high fever (>39.0) is related to severe case on admission and during hospitalization in our analysis.

The radiological abnormalities are a part of diagnosis, all patients have lesion in radiology, and the radiological progression of Covid-19 in severe patient is gradual. The initial radiological lesion are patchy shadows in bilateral or unilateral lower lungs in outer bands, and then progress to 1/3 of both lower lungs, then involve both upper lungs, and eventually develop into “white lung”. In considering of Covid-19 pneumonia progression and 4 or 5 days interval from symptoms onset to diagnosis, a lung CT scan or X-ray routinely in every 3 or 4 days were recommended for the evaluating progression and early diagnosis of Covid-19.

In concert with recent studies in laboratory tests,^7,14,23,28^ lymphopenia, thrombocytopenia and neutrocytosis in severely ill ones suggest the acute mix-infections of bacteria, virus, and even fungi, the activation of circulating immune cell could amplify immune response cascade, inducing both immune cell and cytokine storm, therefore increasing chances of developing septic shock and the organs failure. Interestingly, the decrease of eosinophils is pronounced in severe patients in our study, and eosinophil counts are even lower in some critically ill patients. Despite the fact that use of glucocorticoids would lead to eosinophilia, we still observed this feature in patients without glucocorticoids administration, the previous study also demonstrated that eosinophils percentage related the severity of Covid-19 pneumonia.^28^ The decrease in eosinophil count may be an important predictor of critically ill patients, the subsequent clinical feature analysis should validate the predictability of eosinophil. Abnormalities in organs function and internal disturbance focused on coagulation, liver function and electrolyte system. Increase of D-dimer and decrease serum level of chloride and calcium occurred in critically ill patients, indicating coagulation disorders secondary to inflammatory cascade after severe infection and electrolyte disturbances. The oxygen therapies are of vital importance for the progressive Covid-19, because ARDS is common complication in severe Covid-19 patients, the PaO_2_/FiO_2_ ratio is clinical predictor in both severity and mortality. The PaO2/FiO2 ratio of Cocid-19 patients indicated the effect of oxygenation and the response to current therapies against hypoxemia.

For the critical illness in adults in Hainan, the common reason of severe patients bears a long waiting time for diagnosis and intensive treatments, including a long time from onset to consultation or a long time from the suspect to laboratory-confirmed. The rapid progress in severe patients illustrates the importance of shorten time of each step, so a bundle of consecutive steps including early test, early diagnosis, early isolation, and early treatment for Covid-19 patients might have contributed to a favorable outcome.

Among 160 discharged patients with Covid-19 infection who met the criteria for hospital release of quarantine, 20 of them conversed to positive RT-PCR test results without any clinical symptoms after 3 to34 days after discharge. At present, the cause of the returning positive of RT-PCR still remains unclear, though there were no evidence shows that kind of patients have the infectivity, we still suggest the current standards for hospital discharge should be reevaluated, and the detection and management of discharged patients should be strengthened.

Our study has some limitations. First, though we have included all the patients of Covid-19 in Hainan province, the sample size was still relatively small and the patients were only from Hainan province, it might be that more clinical features related to Covid-19 would not be identified yet. Secondly, some cases had incomplete record of the radiologic and laboratory testing, given the different procedures among different hospitals.

In conclusion, patients infected with Covid-19 have a severe rate of 21.4% and a fatality rate of 3.6% in Hainan province. The elderly people with diabetes, hypertension and CVD are more likely to develop severe cases. Fever is the most common symptom, though it does not occur in all patients before hospital admission. In light of the rapid spread of Covid-19 around the world, and no specialized medication to treat Covid-19, early identification and diagnosis is important for the prevention and treatment of Covid-19 to prevent patients from developing into critical illness.

## Data Availability

Anonymized data will be shared by request from any qualified investigator with proposals approved by the data committee.

## Funding

This study funded by Chinese Academy of Medical Sciences Innovation Fund for Medical Sciences (2019-I2M-5-023), Hainan Provincial Science and Technology Major Project (ZDKJ201804) and Hainan Social Development Fund (ZDYFXGFY2020004). The funders had no role in study design, data collection and analysis, decision to publish, or preparation of the manuscript.

## Acknowledgements

We thank all the hospital staff members for their efforts in collecting the information that was used in this study, and Chinese Academy of Medical Sciences for the support.

## Notes

### Competing Interest Statement

The authors have declared no competing interest.

